# The Yamanashi Multi-omics Cohort (YMoC): study design of a screening-defined longitudinal metabolic-risk cohort with integrated multi-omics and digital phenotyping

**DOI:** 10.64898/2026.08.18.26360529

**Authors:** Go Goto, Daisuke Hanawa, Keita Naito, Qingbo Seiha Wang, Shu Kanai, Momoko Awaji, Hinako Nishikawa, Hideki Yui, Shota Nishitani, Kunio Miyake, Tadao Ooka

**Affiliations:** Department of Orthopaedic Surgery, Faculty of Medicine, University of Yamanashi, Chuo, Yamanashi, Japan; Taomics Inc., Yamanashi, Japan; Department of Health Sciences, Faculty of Medicine, University of Yamanashi, Chuo, Yamanashi, Japan; RIKEN Center for Integrative Medical Sciences, Yokohama, Kanagawa, Japan; Center for Birth Cohort Studies, University of Yamanashi, Chuo, Yamanashi, Japan; Department of Psychiatry and Behavioral Sciences, School of Medicine, Stanford University, CA, USA; Department of Epidemiology, Faculty of Medicine, University of Yamanashi, Chuo, Yamanashi, Japan

**Keywords:** cohort study, multi-omics, digital phenotyping, prediabetes, longitudinal study, Japan

## Abstract

**Background:** Large-scale biobanks have advanced genomic and epidemiologic research, but many rely on infrequent biological sampling and limited digital phenotyping. The Yamanashi Multi-omics Cohort (YMoC) was established to support longitudinal assessment of molecular, clinical, and behavioural changes in a screening-defined cohort of adults at elevated metabolic risk without diagnosed diabetes.

**Methods:** YMoC is a longitudinal cohort of 215 adults aged 30-70 years in Yamanashi Prefecture, Japan, who met prespecified glycaemic eligibility criteria at health check-up, including fasting plasma glucose 100-125 mg/dL (5.6-6.9 mmol/L) and HbA1c <6.5%. Participants underwent three in-person visits over six months. Measurements include 75-g oral glucose tolerance testing with serial sampling, clinical biochemistry, anthropometry, liver elastography, and collection of blood, urine, stool, and saliva for multi-omics profiling. Between visits, participants wore a Fitbit Inspire 3 and completed daily app-based questionnaires using the Taohealth app. Current molecular data include genome-wide single nucleotide polymorphism array genotyping and longitudinal plasma proteomics in a subset.

**Conclusions:** YMoC is designed to evaluate within-person molecular and phenotypic trajectories in a screening-defined metabolic-risk cohort. The cohort provides a dense longitudinal resource linking clinical assessments, biospecimens, omics assays, and digital phenotyping, including analyses of insulin-resistance-related markers such as homeostasis model assessment of insulin resistance (HOMA-IR).

## Introduction

Large-scale biobanks and cohort studies have advanced genomic and epidemiologic research by linking biospecimens with clinical data across large populations.[1–4] Representative resources such as BioBank Japan (BBJ) and UK Biobank have enabled large-scale discovery in complex diseases, but most rely on sparse molecular sampling and limited high-frequency behavioural data.[5–8]

These infrastructures provide breadth of coverage, but they usually measure molecular layers at only one or a few time points. As a result, they are less suited to evaluating within-person temporal dynamics across molecular, clinical, and behavioural domains.[9,10]

Smaller longitudinal multi-omics studies have shown that repeated profiling can reveal dynamic molecular responses and early metabolic change, but such studies have generally involved selected cohorts, limited scale, and little integration with continuous real-world behavioural monitoring.[9–11] Wearable devices and smartphone-based questionnaires can capture high-frequency behavioural and physiological data, but these streams are rarely integrated with repeated multi-omics and standardised clinical testing in a defined metabolic-risk cohort.[12–14]

Plasma proteomics is particularly relevant to early metabolic dysregulation, but most proteomic cohorts still provide only one or two time points per individual, limiting assessment of within-person network change over time.[15–17]

We therefore established the Yamanashi Multi-omics Cohort (YMoC), a longitudinal cohort that recruited adults who applied on the basis of their own routine health-check results. YMoC combines repeated clinical phenotyping, three scheduled plasma proteomics measurements in a selected subset using the Olink Explore HT platform (>5,400 protein targets), biospecimen collection for additional omics layers, and continuous wearable and app-based monitoring in adults meeting prespecified glycaemic eligibility criteria. The cohort is designed to support analyses of within-person molecular and behavioural trajectories in relation to short-term metabolic change, including longitudinal analyses of insulin-resistance-related markers such as homeostasis model assessment of insulin resistance (HOMA-IR).[10,15,16]

In this paper, we describe the rationale, design, and current data availability and structure of YMoC and position it relative to existing biobank and longitudinal multi-omics resources.

## Methods

### Study design and setting

YMoC (Yamanashi Multi-omics Cohort) is a single-site longitudinal cohort study conducted by the Department of Health Sciences, Faculty of Medicine, University of Yamanashi, in collaboration with local health-check providers. Routine health-check results provided by applicants were used for pre-enrolment screening.

The cohort enrolled adults who met prespecified glycaemic eligibility criteria at health check-up, including fasting plasma glucose 100-125 mg/dL (5.6-6.9 mmol/L) and HbA1c <6.5%.

### Eligibility criteria and recruitment

Adults aged 30–70 years who had undergone routine workplace health check-ups or municipally organised health check-ups in fiscal year 2023 or 2024 were invited through recruitment materials to apply on the basis of their own health-check results. The health-check results provided by applicants were used as pre-enrolment screening information for YMoC. Eligibility required fasting plasma glucose 100–125 mg/dL (5.6–6.9 mmol/L), HbA1c <6.5%, no self-reported use of medication for type 2 diabetes, and haemoglobin ≥12.5 g/dL in men or ≥12.0 g/dL in women.

Because eligibility was based on these pre-enrolment health-check values rather than baseline study-visit measurements, some visit-1 fasting plasma glucose or HbA1c values fell outside the screening thresholds.

Recruitment information describing the eligibility criteria was circulated through collaborating organisations in Yamanashi Prefecture. Individuals who considered themselves eligible on the basis of their own health-check results contacted the study team to apply. Recruitment took place between September and October 2024. Before the first visit, the study team mailed participants written study information and specimen kits for stool, urine, and saliva collection.

A total of 215 participants provided written informed consent after an in-person explanation before the baseline visit. Of these, 214 (99.5%) completed the second and third visits over six months; one participant withdrew because of difficulty using the Taohealth app.

### Sample size rationale

The target sample size (n=215) was determined by the practical capacity to deliver repeated oral glucose tolerance tests (OGTTs) and multi-omics assays across three visits.

### Visit schedule

Each participant was scheduled for three in-person study visits over approximately six months. Visit 1 (baseline) occurred in October–November 2024. Visit 2 (3-month follow-up) occurred in January–February 2025. Visit 3 (6-month follow-up) occurred in April–June 2025. The visits spanned autumn, winter, and spring.

Between visits, participants wore a Fitbit Inspire 3 continuously and used the Taohealth smartphone application for daily questionnaires.

The visit structure and between-visit streams are summarised in Fig. 1 and Table 1.

**Fig. 1.**
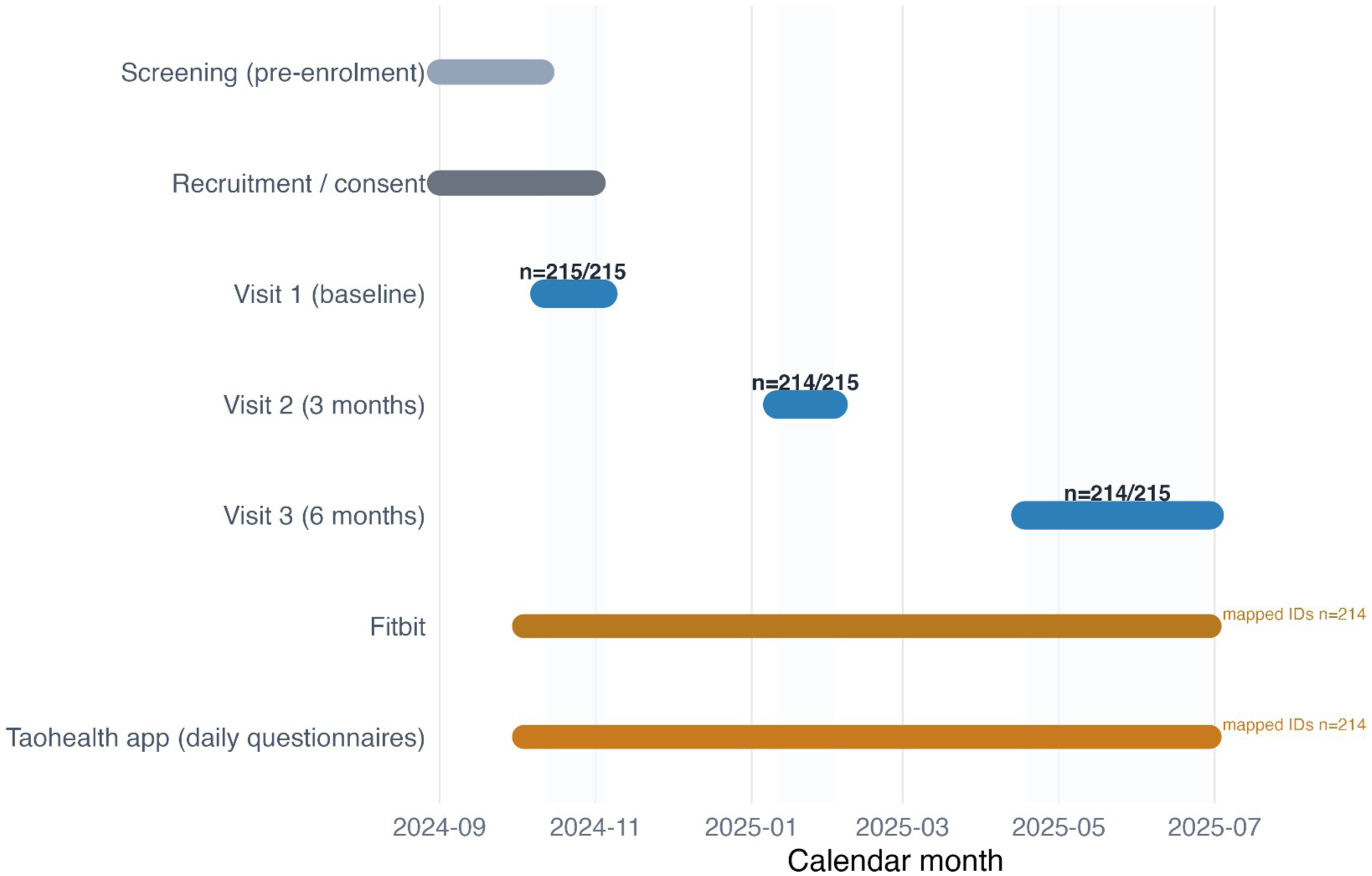
Visit timeline and between-visit digital streams This timeline summarises pre-enrolment screening/recruitment (September-October 2024), three in-person visits (baseline, 3 months, 6 months), and between-visit digital phenotyping windows. Observed clinical visit windows were October-November 2024 (visit 1), January-February 2025 (visit 2), and April-June 2025 (visit 3).

**Table 1.** Schedule of assessments in YMoC (pre-enrolment screening, baseline, 3 months, and 6 months).

| Section | Assessment | Pre-enrolment / screening | Baseline | 3 months | 6 months |
| --- | --- | --- | --- | --- | --- |
| Screening | Eligibility screening (annual health check) | Yes | No | No | No |
| Clinical phenotyping | OGTT (0/30/120 min glucose + insulin) | No | Yes | Yes | Yes |
| Clinical phenotyping | Standard clinical biochemistry / CBC | No | Yes | Yes | Yes |
| Clinical phenotyping | Anthropometry / body composition | No | Yes | Yes | Yes |
| Clinical phenotyping | Liver elastography (FibroScan) | No | Yes | Yes | Yes |
| Biospecimens | Blood, urine, stool, saliva | No | Yes | Yes | Yes |
| Omics | Genome-wide genotyping | No | Yes | No | No |
| Omics | Plasma proteomics (Explore HT subset) | No | Yes | Yes | Yes |
| Questionnaires | General health and lifestyle questionnaire | No | No | Yes | No |
| Questionnaires | Food-frequency questionnaire (FFQ) | No | No | Yes | No |
| Questionnaires | Household and social environment questionnaire | No | No | No | Yes |
| Questionnaires | Exploratory constitution questionnaire | No | No | Yes | Yes |
| Digital phenotyping | Wearable data (Fitbit Inspire 3) | No | Initiated | Continued | Continued to visit 3 |
| Digital phenotyping | Daily app questionnaires (Taohealth) | No | Initiated | Continued | Continued to visit 3 |

**Fig. 1** Visit timeline and between-visit digital streams Fig. 1 presents the three in-person visits (baseline, 3 months, and 6 months), with continuous wearable monitoring and daily app-based questionnaires running between visits.

**Table 1. Schedule of assessments in YMoC (pre-enrolment screening, baseline, 3 months, and 6 months).**

### Clinical assessments at each visit

All visits were conducted in the morning after an overnight fast, following a standardised protocol.

#### Anthropometry and body composition

Height, weight, waist circumference, and body mass index (BMI) were measured. Segmental body composition, including limb and trunk muscle mass, was measured using a bioimpedance analyser (Tanita MC-780).

#### Liver elastography

Liver elastography was assessed using FibroScan-derived liver stiffness and controlled attenuation parameter (CAP). These indices were collected longitudinally across scheduled visits to characterise hepatic status over time.

#### Oral glucose tolerance test (OGTT)

A standard 75-g OGTT was performed with blood sampling at 0, 30 and 120 minutes for plasma glucose and immunoreactive insulin. OGTT categories (normal, borderline, and diabetic patterns) were determined according to Japanese Diabetes Society criteria [18]. Derived indices included HOMA-IR, HOMA-β, insulinogenic index and C-peptide index.

#### Blood tests (non-omics)

Non-omics blood-test items measured across visits are summarised in Table 2.

**Table 2.** Non-omics blood-test item inventory and visit availability.

| Domain | Measured blood items | Visit availability | Notes |
| --- | --- | --- | --- |
| Haematology | WBC, RBC, Hb, Ht, PLT, MCV, MCH, MCHC | V1-V3 | Standard haematology panel |
| OGTT and glycaemic markers | HbA1c; glucose at 0, 30, and 120 min; insulin at 0, 30, and 120 min; fasting C-peptide immunoreactivity | V1-V3 | OGTT-related measures |
| Derived glycaemic indices | HOMA-IR, HOMA-β, insulinogenic index, CPI | V1-V3 | Derived from fasting and OGTT measures |
| Core biochemistry panel | T-P, BUN, UA, creatinine, LDL-C, HDL-C, TG, TBIL, AST, ALT, gamma-GT, cholinesterase, AMY, CK, Na, Cl, K, Fe, LD, ALP | V1-V3 | Core chemistry panel |
| Visit-specific additions | ALB, A/G | V2-V3 | Added from visit 2 onward |
| Visit-specific additions | high-sensitivity CRP, qualitative CRP, quantitative CRP | V3 only | Added at visit 3 |

Non-omics blood-test datasets were curated across all three visits, with item-level availability summarised here; some analytes were introduced from visit 2 or visit 3. Abbreviations: A/G, albumin/globulin ratio; ALB, albumin; ALP, alkaline phosphatase; ALT, alanine aminotransferase; AMY, amylase; AST, aspartate aminotransferase; BUN, blood urea nitrogen; CBC, complete blood count; CK, creatine kinase; Cl, chloride; CPI, C-peptide index; CRP, C-reactive protein; Fe, iron; gamma-GT, gamma-glutamyltransferase; Hb, haemoglobin; HDL-C, high-density lipoprotein cholesterol; HOMA-β , homeostasis model assessment of beta-cell function; HOMA-IR, homeostasis model assessment of insulin resistance; Ht, haematocrit; K, potassium; LD, lactate dehydrogenase; LDL-C, low-density lipoprotein cholesterol; MCH, mean corpuscular haemoglobin; MCHC, mean corpuscular haemoglobin concentration; MCV, mean corpuscular volume; Na, sodium; OGTT, oral glucose tolerance test; PLT, platelet count; RBC, red blood cell count; TBIL, total bilirubin; TG, triglycerides; T-P, total protein; UA, uric acid; WBC, white blood cell count.

#### Biospecimen collection for multi-omics

At each visit, fasting blood (multiple tubes for serum, EDTA plasma, buffy coat and peripheral blood cells) was collected. First-morning urine, stool and saliva brought from home were also collected.

All biospecimens were processed according to standard operating procedures and stored at –80°C in multiple aliquots.

#### Questionnaires

Table 1 summarises the timing of questionnaire administration across visits, whereas the main questionnaire domains are outlined below.

Questionnaires included a general health and lifestyle questionnaire (visit 2), a food-frequency questionnaire (FFQ) (visit 2), and a household and social environment questionnaire (visit 3). Additional items cover psychological factors (including psychological distress, K6), social factors (including social capital), socio-economic variables (including education, income, occupation, and working hours), and lifestyle and medical-history items (including smoking, medical history, and vaccination history).

#### Exploratory constitution questionnaire

At visits 2 and 3, participants completed an exploratory constitution questionnaire, including Ayurveda-based Vata, Pitta, and Kapha items and related behavioural tendencies.

### Multi-omics measurements

The YMoC multi-omics programme is designed to capture dynamic molecular changes across multiple layers in the same individuals.

#### Genome

At baseline, DNA samples from all 215 participants entered the Infinium Asian Screening Array-24 v1.0 single nucleotide polymorphism (SNP)-array genotyping workflow, targeting approximately 700,000 markers, and genotype data processing is ongoing.

#### Longitudinal plasma proteomics

The proteomics subset was selected from participants with visit-1 HbA1c <6.5% and non-diabetic OGTT patterns, including those with borderline or high-normal glycaemic profiles and those with normal glycaemic profiles, after exclusions for visit completion, FibroScan availability, and OGTT timing quality.

Plasma samples were analysed using the Olink Explore HT platform, covering more than 5,400 protein targets, with standardised quality control procedures.

The currently available proteomics dataset comprises 500 sample-level normalised protein expression (NPX) profiles across three visits (visit 1 n=172, visit 2 n=172, visit 3 n=156).

The lower visit-3 participant count reflects the allocation of 16 assay positions to bridging samples across batches in the current NPX release.

Proteomics preprocessing included sample-level QC and run/batch diagnostics, with the bridging samples used to support harmonisation across runs.

#### Additional omics layers

Additional omics layers, including metabolomics, epigenomics, transcriptomics, and gut microbiome profiling, are planned. Biospecimens required for these assays have already been collected according to protocol across scheduled visits.

### Digital phenotyping data

#### Wearable devices (Fitbit Inspire 3)

At baseline, all participants were provided with a Fitbit Inspire 3 device and instructed to wear it continuously for six months. Core wearable measures include heart rate, physical activity, sedentary behaviour, and sleep timing/duration, which are used to quantify day-to-day lifestyle and physiological patterns. Additional vendor-derived summaries, including heart-rate variability (HRV), respiratory rate, oxygen saturation during sleep, skin temperature, sleep-stage estimates, and fitness estimates, are retained when available through the data interface and treated as supplementary wearable-derived variables.

Wearable data are synchronised via Taohealth and stored alongside multi-omics and clinical data. Day-level and intraday Fitbit files are available for activity, steps, calories, zone minutes, heart-rate time series, sleep, HRV (summary and intraday), breathing rate, oxygen saturation, skin temperature, estimated VO2 max (cardio fitness score), and weight.

#### Daily app-based questionnaires (Taohealth)

Taohealth delivered brief daily questionnaires that could be completed in approximately one minute. It captured sleep-related items (including sleep quality and disturbances), dietary and meal-related items (including mealtimes, meal composition, snacking, and intake of sweets, sugar-sweetened beverages, and caffeine), lifestyle items (including alcohol and tobacco use and bowel movements), activity-related items (including step counts and sedentary time, self-reported and synchronised from Fitbit), psychological and social items (including mood, stress, stress-coping strategies, and social-contact-related items), and linked body-metric items (including weight and body fat). The app functioned as a recording and self-monitoring tool and did not provide tailored feedback or behavioural coaching.

Earlier work on smartphone-based digital phenotyping suggests that such passive and active data streams are feasible and acceptable.[12] In the Taohealth daily dataset used for the present analyses, 36,962 daily records are available for 214 participants. Questionnaire summaries are generated using predefined windows.

App-derived variables are defined using prespecified daily response windows and windowed summary features (for example, short look-back summaries before each visit).

### Data management and governance

All data are stored on secure servers with de-identification and role-based access. A unique cohort ID links multi-omics, clinical, wearable, and app data.

Data linkage and curation are summarised in Fig. 2.

**Fig. 2.**
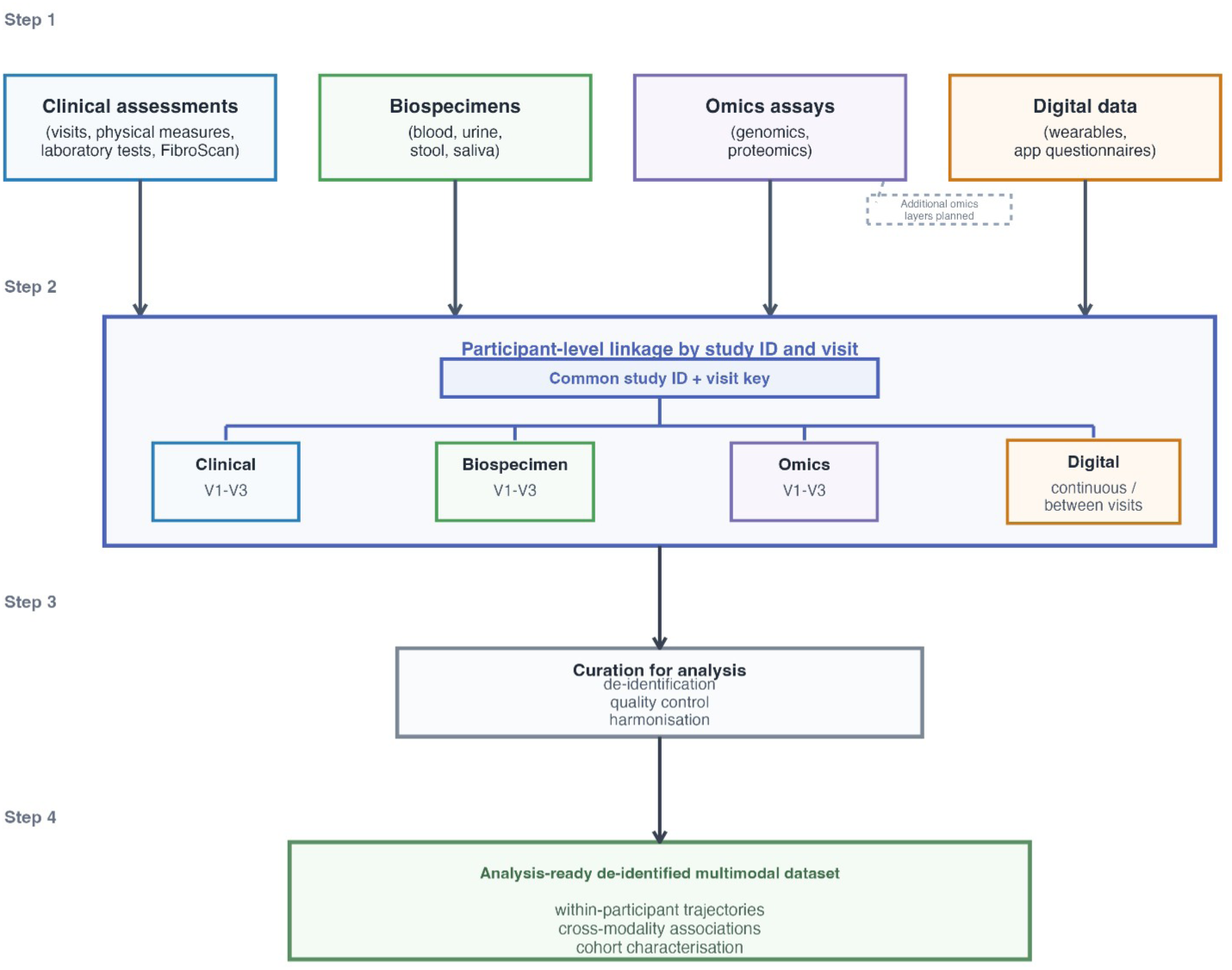
YMoC multimodal data integration workflow Clinical assessments, biospecimens, omics assays, and digital data are linked at the participant level using a common study ID and visit/timepoint information. The linked records are then curated through de-identification, quality control, and harmonisation to generate an analysis-ready de-identified longitudinal multimodal dataset for within-participant trajectory analyses, cross-modality investigations, and cohort characterisation.

### Patient and public involvement

No formal patient or public involvement process was used in the initial study design. During implementation, operational feedback from participants, collaborating organisations, and municipal partners was used to refine participant communication and burden-reduction procedures.

### Ethical considerations

YMoC was approved by the Ethics Committee of the Faculty of Medicine, University of Yamanashi (approval number: CS0052). Participants received an in-person explanation and provided written informed consent covering collection of biospecimens, multi-omics analyses, digital phenotyping, and future approved research use within the scope of consent. Procedures emphasise transparency about data flows and privacy, reflecting concerns raised in the digital phenotyping literature.[12]

Participants may withdraw consent at any time, and handling of previously collected data follows consent scope, governance policy, and applicable regulations. Selected study-visit results are communicated to participants according to the approved protocol and participant consent.

## Results

### Cohort profile

Between September and October 2024, 215 adults meeting the glycaemic eligibility criteria were enrolled and completed baseline assessment. Retention over the six-month protocol was high: 214/215 (99.5%) completed both the 3-month and 6-month visits, and one participant withdrew because they could not continue using the Taohealth app.

**Fig. 3** Participant flow and modality availability in YMoC Participant flow and modality availability are shown in Fig. 3.

**Fig. 3.**
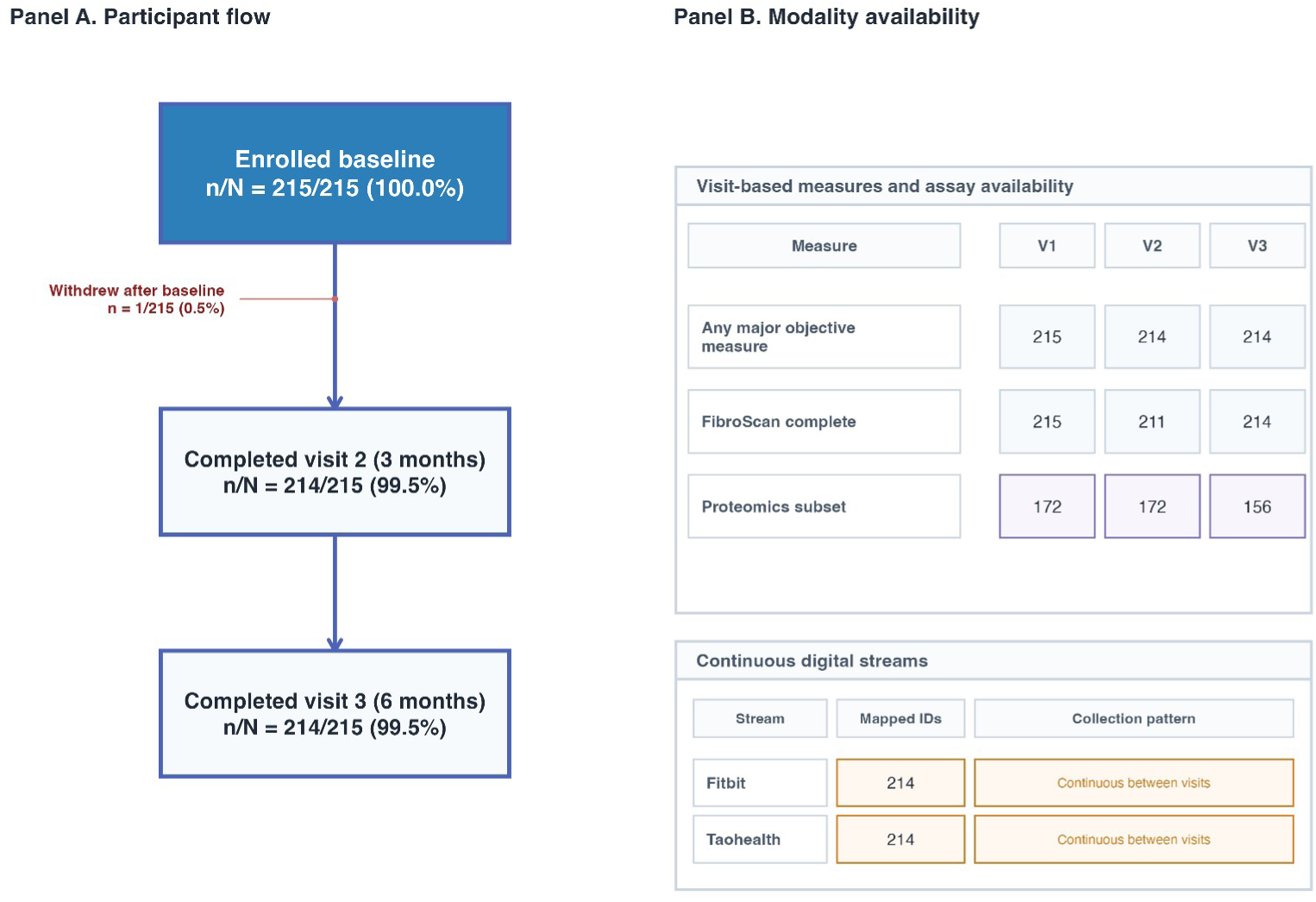
Participant flow and modality availability in YMoC. Panel A reports participant flow with fixed n/N notation: enrolled 215/215, visit-2 completion 214/215, visit-3 completion 214/215, and one withdrawal after baseline due to inability to continue using the Taohealth app. Panel B summarises visit-based measures and assay availability together with participant-level linkage for continuous digital streams.

Baseline cohort characteristics are summarised in Table 3.

**Table 3.** Baseline cohort characteristics in YMoC. Values are n (%) or mean (SD). Glycaemic categories are based on baseline study-visit values; eligibility screening used pre-enrolment health-check values. No missing data were present for the variables shown.

| Domain | Characteristic | Value |
| --- | --- | --- |
| Cohort size | Enrolled participants | 215 |
| Demographics | Sex | Male 131 (60.9%); female 84 (39.1%) |
| Demographics | Age, years | 54.63 (7.76) |
| Anthropometry | BMI, kg/m<sup>2</sup> | 24.76 (3.90) |
| Anthropometry | BMI categories, kg/m<sup>2</sup> | <18.5: 5 (2.3%); 18.5-24.9: 118 (54.9%);<br>25.0-29.9: 70 (32.6%); ≥30.0: 22 (10.2%) |
| Glycaemic markers | Fasting plasma glucose, mg/dL | 101.70 (10.97) |
| Glycaemic markers | Fasting plasma glucose categories, mg/dL | <100: 96 (44.7%); 100-109: 81 (37.7%);<br>110-125: 32 (14.9%); ≥126: 6 (2.8%) |
| Glycaemic markers | HbA1c, % | 5.83 (0.42) |
| Glycaemic markers | HbA1c categories, % | <5.7: 68 (31.6%); 5.7-6.4: 137 (63.7%);<br>≥6.5: 10 (4.7%) |
| Glycaemic markers | Fasting insulin, µU/mL | 6.80 (4.95) |
| Glycaemic markers | HOMA-IR | 1.76 (1.44) |

Table 3 and Fig. 4 use visit-1 measurements, which may differ from the pre-enrolment values used for eligibility screening.

**Fig. 4.**
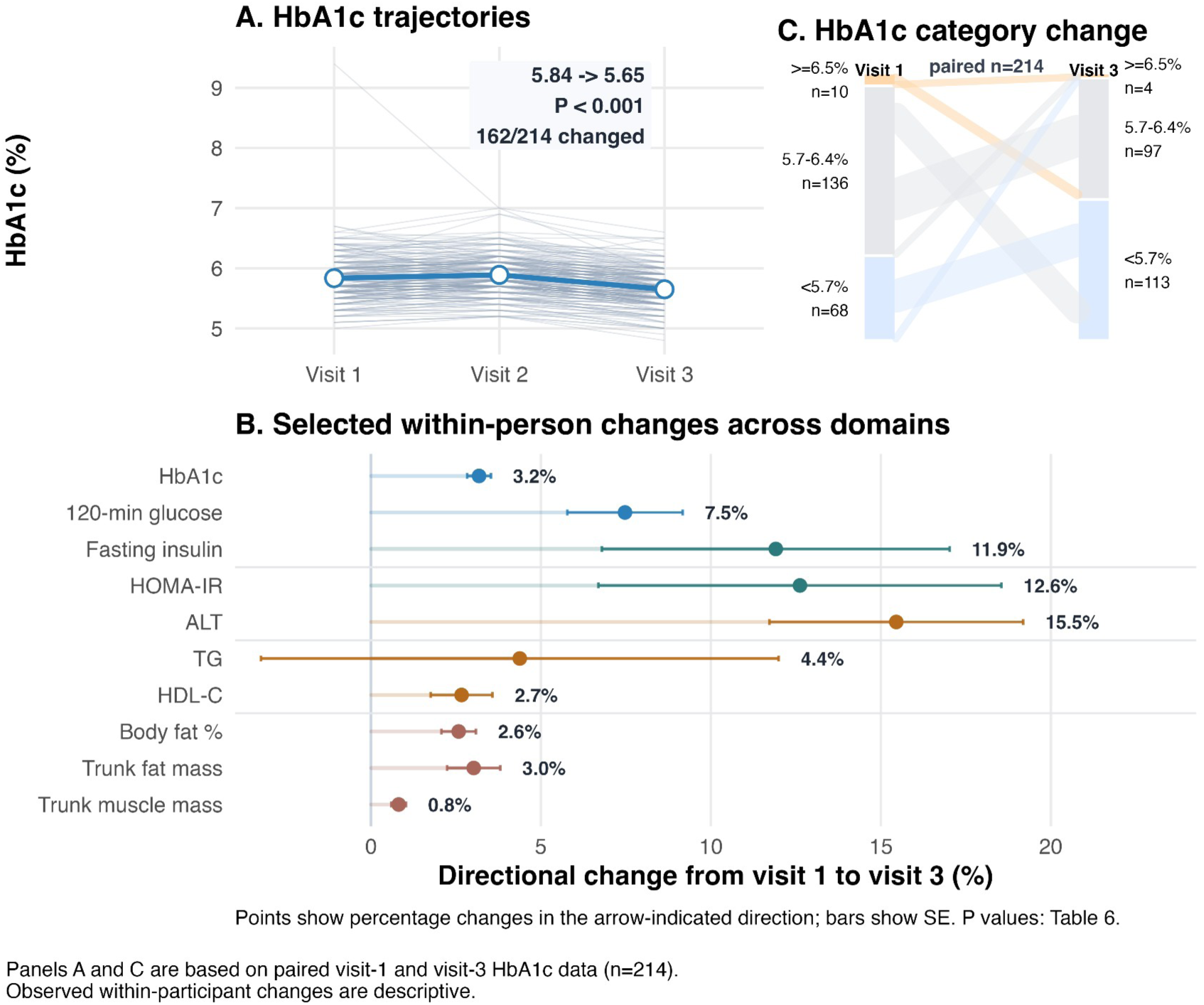
Observed within-person changes in selected markers from visit 1 to visit 3. Panel A shows participant-level HbA1c trajectories across visits with cohort mean values overlaid; the displayed P value is from a paired Wilcoxon signed-rank test comparing visits 1 and 3. Panels A and C are based on paired visit-1 and visit-3 HbA1c data (n=214). Panel B summarises directional change percentages from visit 1 to visit 3 across glycaemic, insulin-related, liver/lipid, and body-composition domains; points indicate percentage changes in the arrow-indicated direction and horizontal bars indicate standard errors. Arrows denote the direction generally considered favourable for descriptive summaries. Panel C shows HbA1c category change from visit 1 to visit 3. Detailed visit means, paired sample sizes, directional change percentages, proportions of participants whose values changed in the indicated direction, and P values for the selected markers are reported in Table 6.

**Table 3. Baseline cohort characteristics in YMoC**.

Data completeness by modality is summarised in Table 4.

**Table 4.** Data completeness across major YMoC modalities.

| Modality / stream | Baseline available N | 3-month available N | 6-month available N | Mapped participant IDs | Collection pattern |
| --- | --- | --- | --- | --- | --- |
| Visit completion | 215/215 (100.0%) | 214/215 (99.5%) | 214/215 (99.5%) | N/A | Visit-based |
| Any objective clinical measure | 215 | 214 | 214 | N/A | Visit-based |
| FibroScan complete metrics | 215 | 211 | 214 | N/A | Visit-based |
| DNA samples entered SNP-array workflow | 215 | Not applicable | Not applicable | N/A | Baseline only |
| Plasma proteomics (subset, current NPX availability) | 172 | 172 | 156 | N/A | Visit-based |
| Wearable stream (Fitbit) | N/A | N/A | N/A | 214 | Continuous between visits |
| App questionnaire stream (Taohealth) | N/A | N/A | N/A | 214 | Continuous between visits |

Abbreviations: N/A, not applicable; NPX, normalised protein expression; SNP, single nucleotide polymorphism.

**Table 4. Data completeness across major YMoC modalities**.

Table 5 summarises currently available datasets and planned resource expansion.

**Table 5.** YMoC data resource overview: current status and planned expansion.

| **Data layer** | **Current status in YMoC** | **Planned expansion** |
| --- | --- | --- |
| Core clinical phenotyping (OGTT, labs, anthropometry, FibroScan) | Three scheduled visits completed for retained cohort (completion n=214 at both V2 and V3) | Harmonised longitudinal tables and data dictionaries |
| Biospecimen inventory | Three-visit collection completed (blood/urine/stool/saliva) | Extended assay-derived variables and sample metadata |
| Genotype data | DNA samples from 215 participants entered the baseline SNP-array workflow | Post-genotyping QC and imputation workflow details |
| Plasma proteomics | Subset NPX available: V1 n=172, V2 n=172, V3 n=156 | Finalised per-visit matrices and QC summaries |
| Wearable and app digital phenotyping | Participant-level digital phenotyping linkage has been completed for Fitbit (n=214) and Taohealth (n=214) | Coverage summaries and derived feature dictionaries |
| Additional omics (metabolomics, epigenomics, transcriptomics, microbiome) | Planned | Stepwise data generation and integration |

**Table 5. YMoC data resource overview: current status and planned expansion.**

### Observed within-person changes in major metabolic and body-composition measures

Descriptive analyses of selected metabolic and body-composition markers were conducted using paired visit-1 and visit-3 data to summarise observed within-person changes during cohort participation.

Mean HbA1c was lower at visit 3 than at visit 1 (5.65% vs 5.84%; paired Wilcoxon signed-rank test, P < 0.001). Similar within-person shifts were observed for selected insulin-related, liver/lipid, and body-composition measures.

Fig. 4 summarises observed within-person changes in selected markers across glycaemic, insulin-related, liver/lipid, and body-composition domains. Table 6 reports the corresponding visit 1 and visit 3 means, directional change percentages, proportions of participants whose values changed in the indicated direction, and P values from paired analyses.

**Table 6.**
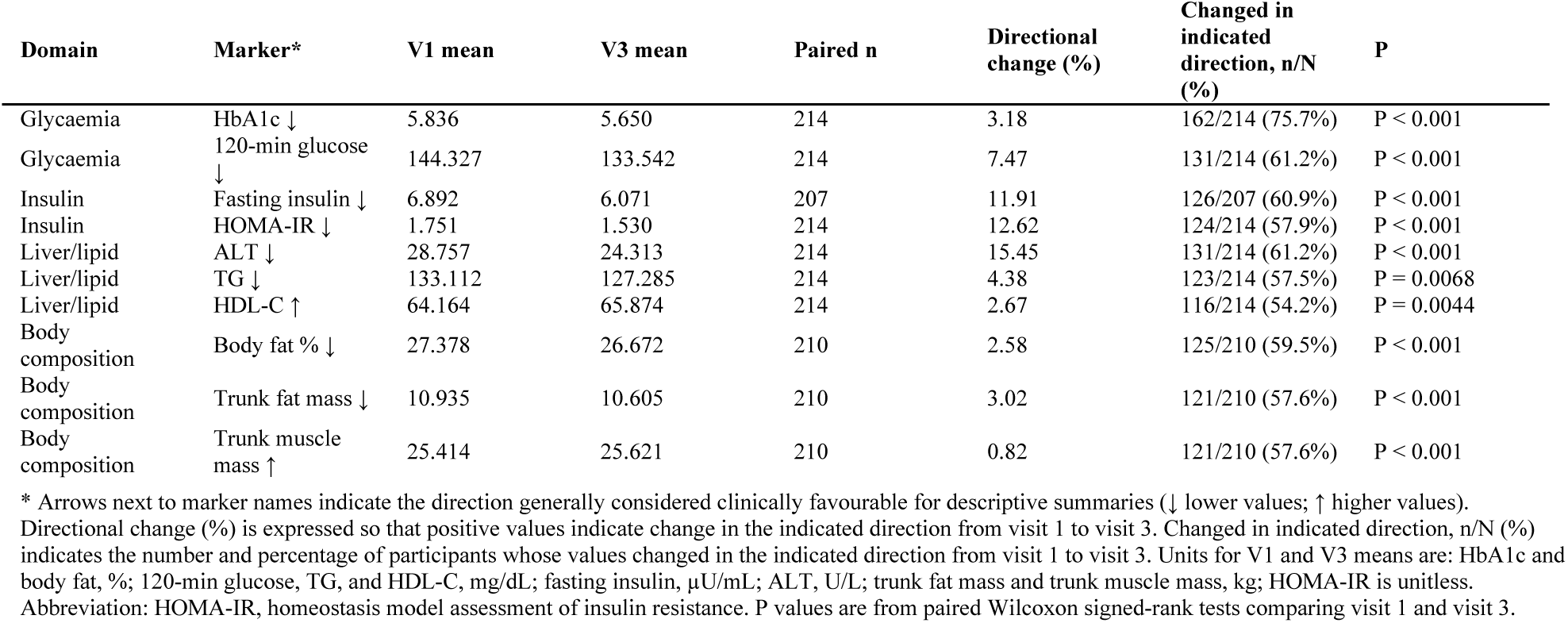
Descriptive summary of observed within-person changes in selected metabolic and body-composition markers.

| Domain | Marker* | V1 mean | V3 mean | Paired n | Directional change (%) | Changed in indicated direction, n/N (%) | P |
| --- | --- | --- | --- | --- | --- | --- | --- |
| Glycaemia | HbA1c ↓ | 5.836 | 5.650 | 214 | 3.18 | 162/214 (75.7%) | P < 0.001 |
| Glycaemia | 120-min glucose ↓ | 144.327 | 133.542 | 214 | 7.47 | 131/214 (61.2%) | P < 0.001 |
| Insulin | Fasting insulin ↓ | 6.892 | 6.071 | 207 | 11.91 | 126/207 (60.9%) | P < 0.001 |
| Insulin | HOMA-IR ↓ | 1.751 | 1.530 | 214 | 12.62 | 124/214 (57.9%) | P < 0.001 |
| Liver/lipid | ALT ↓ | 28.757 | 24.313 | 214 | 15.45 | 131/214 (61.2%) | P < 0.001 |
| Liver/lipid | TG ↓ | 133.112 | 127.285 | 214 | 4.38 | 123/214 (57.5%) | P = 0.0068 |
| Liver/lipid | HDL-C ↑ | 64.164 | 65.874 | 214 | 2.67 | 116/214 (54.2%) | P = 0.0044 |
| Body composition | Body fat % ↓ | 27.378 | 26.672 | 210 | 2.58 | 125/210 (59.5%) | P < 0.001 |
| Body composition | Trunk fat mass ↓ | 10.935 | 10.605 | 210 | 3.02 | 121/210 (57.6%) | P < 0.001 |
| Body composition | Trunk muscle mass ↑ | 25.414 | 25.621 | 210 | 0.82 | 121/210 (57.6%) | P < 0.001 |

\* Arrows next to marker names indicate the direction generally considered clinically favourable for descriptive summaries (↓ lower values; ↑ higher values). Directional change (%) is expressed so that positive values indicate change in the indicated direction from visit 1 to visit 3. Changed in indicated direction, n/N (%) indicates the number and percentage of participants whose values changed in the indicated direction from visit 1 to visit 3. Units for V1 and V3 means are: HbA1c and body fat, %; 120-min glucose, TG, and HDL-C, mg/dL; fasting insulin, $\mu$ U/mL; ALT, U/L; trunk fat mass and trunk muscle mass, kg; HOMA-IR is unitless. Abbreviation: HOMA-IR, homeostasis model assessment of insulin resistance. P values are from paired Wilcoxon signed-rank tests comparing visit 1 and visit 3.

**Fig. 4** Observed within-person changes in selected markers from visit 1 to visit 3

**Table 6. Descriptive summary of observed within-person changes in selected metabolic and body-composition markers.**

## Discussion

### Positioning YMoC relative to existing biobanks and multi-omics cohorts

#### Complementarity with disease-based and population-based biobanks

YMoC adds dense within-person longitudinal multi-omics and daily digital phenotyping over six months in adults recruited using routine health-check results provided by applicants as pre-enrolment screening information. Using these results offered a pragmatic route to recruiting adults who met glycaemic eligibility criteria before overt type 2 diabetes. In contrast to large national biobanks, YMoC prioritises temporal depth over breadth of sample size.[6,7,19,20] This recruitment profile differs from BBJ’s primarily patient-based recruitment and the broader population sample in UK Biobank.[6,7,21]

The YMoC design allows repeated alignment of clinical, molecular, and behavioural data within participants over a short interval.

Taken together, YMoC differs from large population– and disease-based biobanks by prioritising temporal depth, repeated multi-omics, and continuous digital phenotyping within a defined metabolic-risk cohort.

#### Extending longitudinal multi-omics paradigms

Prior longitudinal multi-omics work has shown that dense profiling of individuals can reveal personalised molecular responses to infections, stresses, lifestyle changes, and ageing.[9,10] It can also provide early molecular indicators of metabolic deterioration. These studies have been instrumental in demonstrating feasibility and scientific value but typically involve small numbers of participants, limited integration with wearable data, and ad hoc clinical phenotyping.

YMoC extends this paradigm by anchoring multi-omics in a defined pre-disease glycaemic phenotype based on prespecified eligibility and repeated OGTT-based assessment rather than convenience sampling. It incorporates dense molecular measurements in a prospective cohort with standardised clinical assessments, structured questionnaires, and continuous wearable and app-based data streams, enabling integrated analysis of behavioural, molecular, and clinical trajectories.[9,10,14–16]

### Scientific opportunities enabled by YMoC

The current design supports future analyses of within-person molecular, clinical, and behavioural trajectories, including longitudinal analyses of insulin-resistance-related markers such as HOMA-IR.

#### Digital phenotyping and behavioural signatures

Continuous wearable and app-based data allow quantification of lifestyle and behavioural patterns, including sleep, physical activity, sedentary time, meal timing, diet-related behaviours, stress, and mood. These digital phenotypes can be analysed as time-varying behavioural exposures in relation to short-term metabolic trajectories and longitudinal proteomic changes. [12,14]

#### Proteomic trajectories and metabolic change

Repeated proteomics and OGTTs provide a framework for evaluating within-person proteomic trajectories in relation to short-term metabolic change and for comparing longitudinal signatures with previously reported protein biomarkers.[10,15,16] The broad coverage of the Olink Explore HT platform may also support organ– and system-level interpretation of protein combinations, informed by recent plasma proteomic studies of organ-specific ageing and disease risk.[22,23]

#### Multi-omic integration and pathway modelling

As additional omics layers become available, YMoC will support integrative analyses across genomics, proteomics, metabolomics, transcriptomics, epigenomics, and microbiome-related measures. These analyses are intended to examine how molecular modules relate to short-term metabolic trajectories. The same framework may also support integration of these molecular layers with continuous wearable and app-based data.[9,10]

#### Digital twin-oriented methodological opportunities

YMoC may also provide a useful resource for methodological work that integrates multi-omics, clinical measures, and real-world behavioural data, including digital twin-oriented modelling. [24]

### Strengths and limitations

#### Strengths

##### Temporal depth of multi-omics

Repeated plasma proteomics over three visits in six months, together with biospecimens already collected and secured for additional omics layers, provides an opportunity to study within-person molecular dynamics among participants with screening-defined metabolic risk. This extends cross-sectional proteomics studies of insulin resistance and diabetes risk.[15,16]

##### Integration with high-frequency digital phenotyping

Continuous wearable data and daily app-based questionnaires allow detailed characterisation of lifestyle and behavioural patterns. This extends prior digital phenotyping and digital health research to metabolic health.[12,14]

##### Health check-up-based recruitment of individuals with metabolic risk

Recruitment through health check-ups identifies participants with metabolic risk before overt diabetes. This parallels but extends strategies used in national biobanks such as BBJ, Tohoku Medical Megabank and All of Us.[6,19,20,25]

##### Clinical phenotyping

Standardised OGTTs, liver elastography, body composition and laboratory tests across scheduled visits enhance interpretability and comparability with existing epidemiologic and clinical studies.

##### Low-intervention digital phenotyping context

sRepeated measurements, wearable monitoring, and app-based logging enable YMoC to characterise participant engagement, data completeness, and real-world digital phenotyping patterns alongside clinical and molecular trajectories in a low-intervention cohort setting.

#### Limitations

##### Moderate sample size

The sample size limits large-scale association screening, but YMoC is designed for repeated within-person analyses and intensive multimodal phenotyping. The cohort is planned to expand over time, which may increase opportunities for validation and subgroup analyses.

##### Geographic and ethnic specificity

The cohort is drawn from a single Japanese prefecture and comprises individuals who attend health check-ups. This specificity may limit generalisability, but it also supports detailed linkage with regional health-system data.

##### Six-month intensive follow-up

Although the intensive six-month protocol provides detailed short-term dynamics, it does not yet capture long-term progression to overt diabetes or complications.[19,20,25]

##### Digital participation requirement

Use of a smartphone app and wearable device may exclude individuals with low digital literacy or those unwilling to share digital data, potentially introducing selection bias. Prior digital phenotyping studies report generally good acceptability but also note privacy and equity concerns.[12] Interpretation of wearable– and app-derived measures should account for adherence, missingness, and dropout patterns.

##### Wearable measurement limitations

Several wearable metrics are derived using proprietary algorithms, and data availability and accuracy may vary across metrics and over device firmware updates. However, prior validation studies support acceptable accuracy for the core metrics most relevant to the planned analyses, particularly steps and heart rate.[26–28] Measurement error may nevertheless remain and should be considered when interpreting wearable-derived variables.

##### Interpretation of observed within-person changes

The short-term changes shown in Fig. 4 and Table 6 are descriptive. Because YMoC has no comparator group and eligibility was based on elevated pre-enrolment health-check values, regression to the mean, seasonal variation, and other time-related influences may have contributed to the observed changes.

### Conclusion

YMoC is a longitudinal cohort resource that links repeated clinical phenotyping, multi-omics sampling, and continuous digital phenotyping in adults recruited on the basis of their own routine health-check results and glycaemic eligibility criteria. Its design and current data availability support within-person analyses of short-term molecular, clinical, and behavioural trajectories and provide a foundation for future longitudinal analyses of metabolic health.

## Data Availability

De-identified participant-level data are not currently publicly available because of privacy and ethical constraints. Future external use or deposition in a repository or biobank may be considered through a governance process, subject to additional ethics review and approval.

## Statements and Declarations

### Funding

This work was supported by JSPS KAKENHI Grant Number JP24K02856, the JST FOREST Program under Grant Number JPMJFR2155, and the Japan Agency for Medical Research and Development (AMED) under Grant Number JP23le0110028.

### Ethics approval and consent to participate

The study was approved by the institutional ethics committee of the University of Yamanashi (approval number: CS0052). All participants received an in-person explanation and provided written informed consent before enrolment.

### Consent to publish

Not applicable.

### Competing interests

Go Goto is employed by and holds shares in Taomics Inc., which owns the Taohealth application used in this study. Tadao Ooka is a founder and shareholder of Taomics Inc. QSW is a full-time employee of Calico Life Sciences. QSW’s involvement in the present study was independent of this affiliation. The remaining authors declare that they have no relevant financial or non-financial interests to disclose.

### Author contributions

Tadao Ooka contributed to study conception and design. Tadao Ooka and Daisuke Hanawa contributed to participant recruitment, data collection, and study coordination. Shu Kanai, Momoko Awaji, and Hinako Nishikawa contributed to participant coordination and data collection during study visits. Shota Nishitani and Kunio Miyake contributed to data collection and biospecimen processing. Hideki Yui provided advice on ethical aspects of the study. Go Goto and Keita Naito contributed to data curation. Go Goto contributed to statistical analysis, figure and table preparation, and drafting of the manuscript. Qingbo Seiha Wang contributed to interpretation of the data and critical revision of the manuscript. All authors critically reviewed the manuscript and approved the final version.

## Acknowledgements

We thank all participants, collaborating health-check providers, and municipal and institutional partners who supported recruitment, follow-up, and data collection.

Generative artificial intelligence tools (ChatGPT and Codex; OpenAI) were used to assist with language editing and the preparation of revision proposals. All AI-assisted outputs were critically reviewed and revised by the authors, who take full responsibility for the final manuscript.

